# Cataract in the human lens: a systematic review of proteomic studies

**DOI:** 10.1101/19009035

**Authors:** Christina Karakosta, Argyrios Tzamalis, Michalis Aivaliotis, Ioannis Tsinopoulos

## Abstract

**Background/Aim:** The aim of this systematic review is to identify all the available data on human lens proteomics with a critical role to age-related cataract formation in order to elucidate the physiopathology of the aging lens.

**Methods:** We searched on Medline and Cochrane databases. The search generated 328 manuscripts. We included nine original proteomic studies that investigated human cataractous lenses.

**Results:** Deamidation was the major age-related post-translational modification. There was a significant increase in the amount of αA-crystallin D-isoAsp58 present at all ages, while an increase in the extent of Trp oxidation was apparent in cataract lenses when compared to aged normal lenses. During aging, enzymes with oxidized cysteine at critical sites included GAPDH, glutathione synthase, aldehyde dehydrogenase, sorbitol dehydrogenase, and PARK7.

**Conclusion:** D-isoAsp in αA crystallin could be associated with the development of age-related cataract in human, by contributing to the denaturation of a crystallin, and decreasing its ability to act as a chaperone. Oxidation of Trp may be associated with nuclear cataract formation in man, while the role of oxidant stress in age-related cataract formation is dominant.

**Synopsis:** The oxidative stress and the post-translational modification of deamidation in lens crystallins seem to play a significant role in the formation of age-related cataract in human.

## Background

Cataract is an opacification of normally clear crystalline lens that may cause a decrease in vision or even blindness if left untreated (1). Cataract is responsible for the 33% of blindness worldwide (2–4). Age is the most important aetiological factor of cataract, due to the cumulative damage of environmental insults on proteins and cells of the crystalline lens (5,6,7).

The lens fibers have a high protein content (∼34%) and relatively low water content (∼65%). The crystallins are water-soluble structural proteins of the lens fibers. Human crystallins are classified into three main types: alpha, beta and gamma crystallins. Beta and gamma crystallins are similar in sequence and structure, so they are grouped as a protein superfamily, the βγ-crystallins. The α-crystallin family and βγ-crystallins represent the majority of proteins in the human lens fibers (8).

The crystallins need an antioxidant environment in order to remain soluble and maintain the transparency of the lens (6,9). In order to elucidate the complex factors involved in cataractogenesis, it is necessary to understand the dynamics of this protein superfamily and proteomics research is an ideal way to do so. With the progress in proteomic analysis, proteomic approaches have been applied to identify proteins in various physiological or pathological conditions in the crystalline lens and analyze the proteome of cataractous or normal lenses. Moreover, various investigations on different aspects of the lens proteomics of different species have been carried out using a wide variety of proteomic techniques (10).

In particular, many studies were focused on lens proteomics of various species, such as mice. In 2002, Ueda Y. et al. suggested that partial degradation of α- and β -crystallins and increased acidity of γ-crystallins may cause insolubilisation during aging and they created 2-DE proteome maps of mouse lens proteins in order to be compared with maps of lens proteins of mice with cataracts and facilitate the identification of cataract-specific modifications (11). In 2006, Hoehenwarter W. et al. published a review of eye lens proteomics that summarized crystallin modifications in the lens detected by proteomics techniques (9).

In 2013, the Human Eye Proteome Project initiated with a principal goal the characterization of the proteome of the human eye. In 2013 there were 4,842 non-redundant proteins identified in the human eye. In 2018, an updated resource of 9,782 non-redundant proteins in the human eye was reported. Regarding the human lens, the number of known proteins in the human lens and zonule system was increased from 273 in 2013 to 1,117 known proteins to date (12). Although considerable advances have been made to clarify the pathophysiology of age-related cataract formation, additional high-quality data focused on the human lens need to be exported using proteomics. Thereby, it is necessary to collect and summarise all proteomic researches to date that provide data on cataractous human lens compared to a healthy human lens in order to elucidate the complex factors involved in cataractogenesis and suggest possible prevention or even new non-invasive treatment of age-related cataract.

## Aim

The present systematic review aims to identify all the available data on human lens proteomics with a critical role to age-related cataract formation in order to elucidate the different physiopathological states that lead to the aging of the crystalline lens and thus, cataractogenesis.

## Methods

A systematic review was conducted according to the Preferred Reporting Items for Systematic Reviews and Meta-Analyses (PRISMA) guidelines (13).

### Eligibility criteria

In this systematic review were included English full-text peer-reviewed articles that used proteomics methodologies to study in vivo or in vitro human lens proteins and define possible pathways for age-related cataract formation.

In particular, studies were selected according to the criteria outlined below.

Type of studies: original English studies that used proteomic analysis were included. Regarding geographical location, publication status or year of publication there were no limitations. Reviews and studies published in non-English language were excluded.

Type of participants: Age-related cataractous human lenses were the object under investigation of the included studies. Studies that set cataractous lenses of other species as the object under investigation were excluded.

Type of intervention: Mass spectrometry-based proteomic analysis, *in vivo* or *in vitro*, was used in the included studies. For the separation and isolation of proteins electrophoresis or other methods were used in the included studies.

Type of outcome: Aging of the crystalline lens and cataract formation was the outcome of interest in the proteomic analysis. Articles that studied other types of cataract except for the age-related one will be excluded.

The eligibility criteria are summarized in Table 1.

**Table 1.**
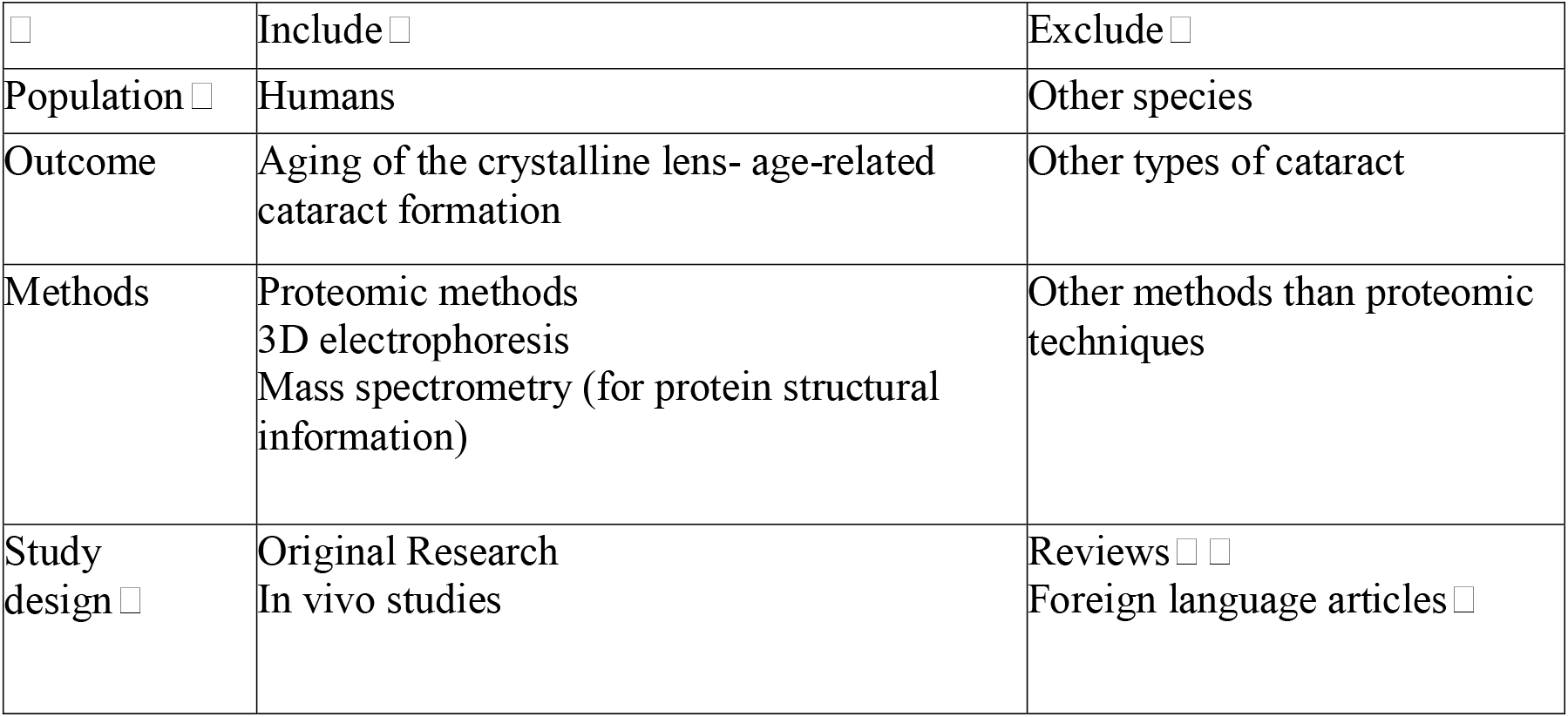
Eligibility criteria

| ☐ | Include ☐ | Exclude ☐ |
| --- | --- | --- |
| Population ☐ | Humans | Other species |
| Outcome | Aging of the crystalline lens- age-related cataract formation | Other types of cataract |
| Methods | Proteomic methods<br>3D electrophoresis<br>Mass spectrometry (for protein structural information) | Other methods than proteomic techniques |
| Study design ☐ | Original Research<br>In vivo studies | Reviews ☐ ☐<br>Foreign language articles ☐ |

### Information sources

Searching electronic databases identified all the available studies. Primarily, two major databases were used for the purposes of this review, PubMed/MEDLINE and Cochrane Database of Systematic Reviews. Further search on PROSPERO for on-going systematic reviews was conducted. Besides, we hand-searched contents pages of Journal of Vision and Journal of Investigative ophthalmology & visual science. In order to ensure literature completeness, we screened additionally the reference lists of the studies that were included in our search. The literature search in the present systematic review was restricted to the English language and human subjects. References identified were imported in Mendeley Desktop software for deduplication.

### Search strategy

Two reviewers independently screened the titles and the abstracts against the eligibility criteria. Our search strategy was based on proteomics and the biological context of interest (human crystalline lens). Our literature search strategies were developed using medical subject headings (MeSH) and free-text terms related to proteomics and crystalline lens. The “English” filter was applied via the Filter section.

We used the following search terms in all databases: proteomics; proteome; “proteom*”;

“proteomic analysis”; “proteomic study”; “lens, crystalline”; “Anterior Capsule of the Lens”; “Posterior Capsule of the Lens”; “Lens Nucleus, Crystalline”; “Lens Cortex, Crystalline”; “Lens Capsule, Crystalline”; “Crystallins”; “Cataract” and “Cataract, Age-Related Nuclear”. The detailed research strategy can be found in Appendix 1.

### Study records

#### Data management

Literature search results were uploaded to Covidence™. Our team members developed screening questions based on the eligibility criteria. Citation abstracts and full-text articles were uploaded to Covidence™ based on those screening questions.

#### Study Selection process

Two principal reviewers, independently working, screened the titles and abstracts of the results retrieved from our search strategy. Full-text forms were obtained for the potentially eligible studies or in case of any uncertainty. Then, the two reviewers assessed the full-text forms and chose those that met the predefined inclusion criteria. In order to avoid data duplication, multiple publications from the same research group were not extracted (if reporting the same methodology and the same results); instead, the paper that provided the most robust evidence was chosen as the primary evidence source.

Any potential disagreements were resolved through discussion with a senior reviewer. All the reasons for excluding studies were recorded. Neither of the principal reviewers nor the senior reviewer was blind to the journal titles or the study authors or institutions.

#### Data collection process

The two principal reviewers using a predefined standardized form for data extraction accomplished data collection process. The first reviewer extracted the data from the included studies and the second one checked the extracted data. Reviewers resolved any disagreements by discussion; if no agreement could be reached a discussion with a senior reviewer was planned. Study authors were contacted if any uncertainty was present.

### Data items

Information was extracted from each study on:

- Study identification characteristics (authors, year of publication, trial registration code, country of origin, corresponding author’s detail)
- Study Characteristics (the type of study: proteomic analysis)
- Type of methods used for separation and isolation of proteins (Electrophoresis or other methods)
- Type of methods used for the obtainment of protein identification, structural characterization and relative quantitation (Mass spectrometry)
- Results (any of the following: identified proteins, protein structural
- information, post-translational modifications, protein expression, protein function, protein localization, protein-protein interactions)
- Role of the identified protein in aging of the crystalline lens and cataract formation

### Assessment of risk of bias

In order to assess the possible risk of bias for each eligible study while collecting data, we created a risk of bias tool based on several questions. The assessment tool covered the following domains: number of subjects, compliance with the manufacturer’ instructions while applying the proteomic techniques, repetition of the experiment. The procedures that took place in each study were described, for each domain of the tool. Studies were rated as of ‘high risk,’ ‘low risk,’ or ‘unclear risk.’ Two independent reviewers conducted the assessment of the risk of bias, and any disagreement was solved by discussion and by consulting a senior reviewer.

### Synthesis of results

A narrative summary of all the included studies was conducted. In case of missing data, we intended to contact the original authors of the study. Results were presented alongside overall judgments for concerns regarding the validity and imprecision of the result.

## Results

### Study selection

A total of nine studies were identified for inclusion in our systematic review. Our search provided a total of 328 citations. In particular, the search of Medline and Cochrane databases provided a total of 300 citations. After adjusting for duplicates still, 300 citations remained. Of these, 134 studies were discarded because they were irrelevant to our subject. After reviewing the abstracts of the rest 166 studies it appeared that 103 papers did not meet the inclusion criteria. The full texts of the rest 63 citations were examined in more detail. After checking the references of located, relevant papers and searching for studies that have cited these papers, 28 additional studies were identified. It appeared that further 82 studies did not meet the inclusion criteria, as described. Thus, nine studies met the inclusion criteria and were included in our systematic review. No unpublished relevant studies were obtained. The flow diagram of the study selection is shown in figure 1. The reasons for exclusion of the 237 papers can be found in table 2. The reasons for exclusion of 82 references after reviewing their full text forms can be found in table 3. The full list of 328 citations can be found in Appendix 2. The exclusion reasons for each citation and the full list of 91 articles that were studied in more detail, can be found in Appendix 3. The full list of articles excluded after reviewing their full text forms, including exclusion reasons for each citation, can be found in Appendix 4.

**Table 2.** Reasons for exclusion of 237 references identified in title and abstract screening for inclusion

| Reason | Number of papers excluded on this basis |
| --- | --- |
| Irrelevant studies | 134 |
| Wrong study design (not an original proteomic study) | 12 |
| Wrong patient population (non-human lens) | 43 |
| Wrong outcome (not age-related cataract) | 48 |
| Total | 237 |

**Table 3.** Reasons for exclusion of 82 references after reviewing their full text

| Reason | Number of papers excluded on this basis |
| --- | --- |
| Wrong study design (not an original proteomic study) | 26 |
| Wrong patient population (non-human lens) | 8 |
| Wrong outcome (not age-related cataract) | 41 |
| Method limitation | 7 |
| Total | 82 |

**Figure 1.**
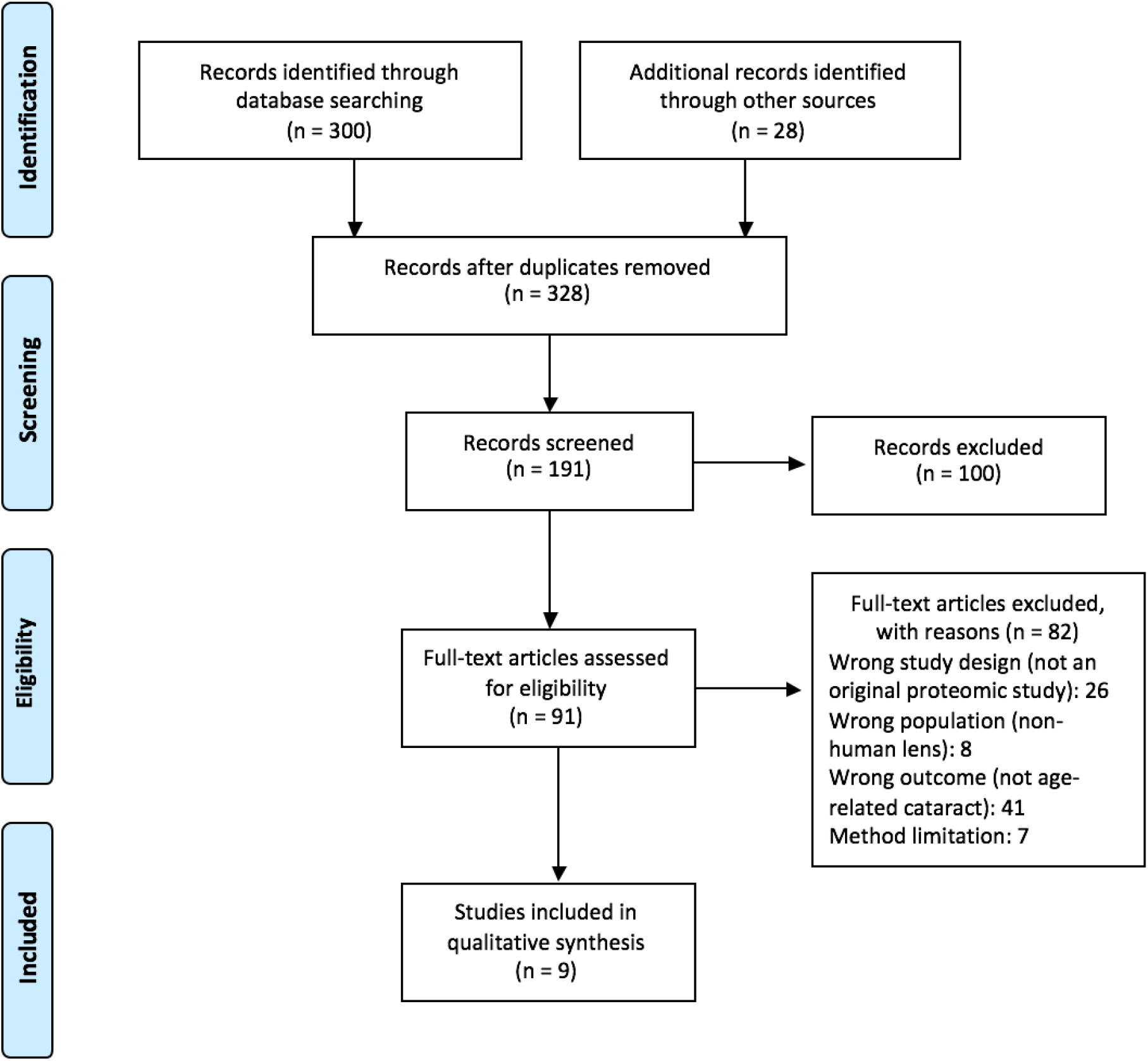
Flow diagram of the study selection

### Study characteristics

The included studies were published between 2005 and 2017. Of those, three were conducted in U.S.A and six in Australia.

Methods: All nine studies finally selected for the review were proteomic analysis studies published in English. All of them used Mass Spectrometry.

Participants: The included studies involved human lenses.

Intervention: They performed proteomic analysis to compare normal human lenses to cataractous human lenses.

Outcomes: The included studies obtained information on proteins with a key role in the aging of the crystalline lens. In all studies, the primary outcome assessed was this information, which was either identified proteins, or protein structural characteristics, or post-translational modifications, or protein expression, or protein function, or protein localization, or protein-protein interactions.

### Risk of bias within studies

The conditions in basic science studies could be completely controlled, in contrast to the clinical studies. As a result, the risk of bias is minimum in basic science studies. However, we created a risk of bias tool to assess the included studies.

The results can be found in table 4.

**Table 4.** Risk of bias within studies

| Studies | Number of subjects | Compliance with the manufacturer' instructions | Risk of bias |
| --- | --- | --- | --- |
| Wilmarth et al. (2005) | 4 | yes | low |
| Hains P.G. and Truscott R.J. (2007) | 6 | yes | low |
| Hains P.G. and Truscott R.J. (2008) | 10 | yes | low |
| Hains P.G. and Truscott R.J. (2010) | 10 | yes | low |
| Hooi et al. (April 2012) | Unclear (>4) | yes | low |
| Hooi et al. (May 2012) | Unclear (>4) | yes | low |
| Hooi et al. (November 2012) | Unclear (>4) | yes | low |
| Fan et al. (2015) | 8 | yes | low |
| Wang et al. (2017) | 6 | yes | low |

### Results of individual studies

All nine studies revealed precious information regarding lens aging and cataract formation. A graphical illustration of the results can be found in figure 2.

**Figure 2.** Graphical illustration of results

Wilmarth et al. (2005) (14) created a comprehensive map of post-translational modifications (PTM) in human lens, analyzing three aged lenses, two of which had moderate cataract, and one young control lens. They identified 155 age-related crystalline PTM sites, including 78 previously unreported sites. Based on their results, large numbers of asparagine and glutamine sites were found to be significantly more deamidated in the water-insoluble proteins of aged lenses and deamidation was found to be the major age-related PTM. They also confirmed the far greater extent of S-methylation known in lens, while there was no methylation in the 3-day-old lens. They identified specific Carboxymethyl lysine (CML) and carboxyethyl lysine (CEL) sites in lens crystallins, which are advanced glycation end products and have been suggested as general markers of advanced age and oxidation, although these modifications were far less abundant than deamidation or methylation.

Hains P.G. and Truscott R.J. (2007) (15) compared PTMs present in young, aged, and cataractous lenses, analysing a total of six human lenses, two young lenses (34, 35 years old), two aged (72, 77 years old), and two early-stage nuclear cataract (70, 80 years old). The researchers found that the proteins from all lenses contained numerous sites of deamidation, oxidation, Cys-methylation, and acetylation. Deamidation was the most common PTM. An increase in the extent of Trp oxidation was apparent in cataract lenses when compared to aged normal lenses. Methylation of Cys in lens proteins was also reported (five Cys residues were detected as being modified in the young lens group, four in the old normal lenses, and three in the cataract lenses). A new phosphorylation site, in γS-crystallin (S89) was identified in one of the young lenses, while, no phosphorylated Ser residues were detected in αB-crystallin from the cataract lenses, whereas up to four were observed in the young and aged normal lens samples. Οne new βB1-crystallin truncation was identified in both the aged normal and cataract lenses. No Arg+55 Da modifications were detected in the cataract lenses.

Finally, Carboxymethyl lysine was detected in all lens groups, two in young, and one each in aged and cataract lenses.

Hains P.G. and Truscott R.J. (2008) (16) examined the lens proteome using foetal (two sets), aged normal (three, 59, 65 and 80 year old), and advanced nuclear cataract lenses (three 60, 65 and 80 year old). They identified 231 proteins across all lens samples, of which 183 in foetal, 103 in adult normal and 105 in cataractous lenses. Of those, 60 proteins were found to be common to all lens, while 22 previously unreported proteins were identified in normal aged and 29 in cataract lenses. They stated as well that for most of the proteins presented, the relative abundance of foetal lens proteins was greater than either of the other lens groups, with the exception of phakinin (BFSP2_HUMAN), protein DJ-1 (PARK7_HUMAN) and phosphoglycerate kinase 1 (PGK1_HUMAN), where both cataract and aged normal lenses had greater relative abundance.

In addition to this, they reported that several proteins had a high abundance in the foetal lens group but a much reduced level in both adult lens groups, including betaine– homocysteine S-methyltransferase 1 (BHMT1_HUMAN), bA2-crystallin (CRBA2_HUMAN), bB3-crystallin (CRBB3_HUMAN), gC-crystallin (CRGC_HUMAN), glycer-aldehyde-3-phosphate dehydrogenase (G3P_HUMAN), per oxiredoxin-6 (PRDX6_HUMAN) and vimentin (VIME_HUMAN).

Hains P.G. and Truscott R.J. (2010) (17) characterized the sites of deamidation in all the major crystallins in the human lens using two sets of fetal lenses, each containing two lenses (set 1, 2 lenses, 17 weeks’ gestation; set 2, 1 lens, 17.5 weeks’ gestation plus 1 lens, 19 weeks’ gestation), three normal (59, 65, 80 years) lenses, and three nuclear cataractous lenses (60, 65, 80 years).

Firstly, they stated that the number of deamidated residues varies considerably between the various crystallins. In βB2- and γC-crystallin, only one residue was found to be deamidated in the aged normal lenses. By contrast, nine sites of deamidation were found in βB1-, eight in βA4-, five in γ-S, and six in βA1/A3-crystallin.

Moreover, the researchers reported that the extent of deamidation varied substantially. Some crystallins from older lenses contained sites that were almost totally deamidated. These included βA1/A3-, βA4-, γS-, and γD-crystallins. In βB2-crystallins, the most highly deamidated residue in older normal lenses was Gln146, and the extent of deamidation was approximately 3%. In αA-crystallin, the most deamidated sites were Asn123 and Gln126, and the extent of deamidation was approximately 10%.

They also observed that the sites of deamidation were often localized to discrete regions. In αA-crystallin, deamidation was found only in the putative C-terminal domain. In βA1/A3-crystallin, the most substantial deamidation was found at three sites contained within a region spanning residues 103 to 133. In γC-crystallin, the only site of deamidation was localized to the peptide bridging the two domains. In most cases, the most highly deamidated sites were those of Asn residues. Finally, in their study, some sites (αA, Q147; αB, N78, Q108; βΑ4, Q64, N82; βΒ1, Ν67, Ν69; γC, N24; γD, N160; γS, N14, Q16, N76) showed increased levels of deamidation in ARNC cataractous lenses.

Hooi et al. (April 2012) (18) studied age-dependent changes in one crystallin representative from each of the two major lens protein groups (α and β/γ crystallin families). Τhe first was an Asp residue in αA crystallin (Asp 151), because it had been shown to be significantly racemised in old human lenses and the second was an Asn residue in γS crystalline (Asn 76) because it was found to be almost twice as deamidated in 60- to 70-year-old cataract lenses as in normal controls. The authors examined lenses across the age range and compared the extent of modification at both sites. For αA-crystallin, there was no significant difference in Asp 151 racemization between cataract and normal lenses. By contrast, in γS-crystallin the degree of conversion of Asn 76 to isoAsp in cataract lenses was approximately double that of normal lenses at every age.

Hooi M.Y. et al. (May 2012) (19) characterized a site of glutamine (Gln) deamidation in γS-crystallin and monitored its rate of conversion to Glu92 as a function of time and compared this with deamidation of Gln170 and Glns in b-sheet regions of the protein. They showed that there was a linear increase in the proportion of deamidation until teenage years, while there was no further increase for the next 50 years. They found no significant difference in the extent of Gln92 and Gln170 deamidation between cataract and age-matched normal lenses.

Hooi et al. (November 2012) (20) investigated the racemisation of Asp 58 in the most abundant structural protein in the human lens, aA-crystallin, comparing normal and cataractous human lenses. They stated that there was a significant increase in the amount of D-isoAsp58 present at all ages. This was most evident during the early years, when age-related cataract is first observed (the average value for D-isoAsp in cataract lenses was almost 26 % compared to normal lenses, almost 12 %).

Fan et al. (2015) (21) created the LEGSKO mouse in which lenticular GSH was lowered by knocking out the γ-glutamyl cysteine ligase subunit Gclc, in order to mimic the oxidative process and formation of protein disulfides linked to low concentrations of glutathione (GSH) in the nucleus of the human lens. These LEGSKO mice developed nuclear cataract by about 9 months. In order to test whether sites of disulfide bond formation are similar in mouse and human age-related cataract, the researchers performed the first comparative analysis of the cataract prone LEGSKO mouse and human aging and cataractous lens crystallin disulfidome, and compared the results with the disulfidome from mouse lens homogenate oxidized in vitro with H2O2 as a model of crystallin aggregation and opacification. They used as control 3 young normal human lens nuclei (at 15, 7, and 3 years of age), and 3 old normal human lens nuclei (at 74, 72, and 68 years of age). They used as well 9 cataractous lenses, 3 in each category of grade II-V of Lens Opacities Classification System III. The age of grade II group was 64, 67, and 60 years old, the grade III was 60, 59, and 69 years old, the grade IV was 56, 63, and 77 years old, and the grade V was 75, 80, and 78 years old.

Fan et al. first studied which proteins are associated with intra-versus intermolecular disulfide-bond in old normal versus young human lens, and they determined how these compared with those detected in oxidatively stressed, precataractous LEGSKO mouse lens. Then they studied how crystallin disulfides found in human cataracts compare with those detected in glutathione-depleted LEGSKO mouse cataracts and mouse lens proteins undergoing in vitro oxidation.

Based on their findings, there was evidence of age-related shift from intra- to intermolecular disulfide formation in human and LEGSKO Mouse Lens Crystallins, since over 25 intramolecular disulfide bond protein spots and less than six intermolecular disulfide protein spots were identified in 4 year old lens, whereas in 67 year old human lens, less than four intramolecular disulphide and more than 20 intermolecular disulfide proteins spots were identified, in combination with a similar shift in spot pattern in the old versus young WT mouse and with the fact that the intra- to intermolecular disulfide shift had already occurred at 5mos in the precataractous, GSH suppressed, LEGSKO mouse lens when compared with the 5 months old WT lens.

Furthermore, 4 types of cysteine residues were identified based on their susceptibility to oxidation with age and as a function of presence and severity of cataract graded from II to V. The first includes cysteine residues that are mildly oxidized with age and not further oxidized in cataracts (Cys67 in βB2, Cys19 in γD, Cys109 in γC, Cys99 in βA4 and Cys185 of βA1/3); the second includes cysteine residues with mild oxidation in old normal lenses but massive increase in cataracts regardless of cataract severity (Cys42 in γD, Cys52 and Cys170 in βA1/3, Cys38 in βB2 and Cys83 in γS); the third includes cysteine residues with oxidation in old normal lenses and further increase in cataract lenses (Cys33 in βA4, Cys80 in βB1, and Cys23 in γC), and the fourth includes cysteine residues that are barely oxidized with age but oxidized in cataracts, and also associated with cataract severity (Cys154 in γC, Cys117 in βA1/3, Cys115, and Cys130 in γS, Cys33 in γD, Cys42 and Cys78/ Cys79 in γC).

Wang et al. (2017) (22) extented their previous study (Fan et al. 2015) in order to identify all noncrystallin disulfides that are common to human and LEGSKO mouse cataracts and in their in vitro oxidation, the sites and patterns of these cysteine residues that are oxidized during aging and oxidation, and the potential impact on lens biology based on their molecular functions. Their results showed that enzymes with oxidized cysteine at critical sites included GAPDH (hGAPDH, Cys247), glutathione synthase (hGSS, Cys294), aldehyde dehydrogenase (hALDH1A1, Cys126 and Cys186), sorbitol dehydrogenase (hSORD, Cys140, Cys165, and Cys179), and PARK7 (hPARK7, Cys46 and Cys53). They stated that extensive oxidation was also present in lens-specific intermediate filament proteins, such as BFSP1 and BFSP12 (hBFSP1 and hBFSP12, Cys167, Cys65, and Cys326), vimentin (mVim, Cys328), cytokeratins, the majority of which were positively associated with degrees of cataract severity (Cys51, Cys77, and Cys474 in keratin 6A; Cys42 and Cys489 in keratin 2; Cys25, Cys66, and Cys427 in keratin 10; Cys55 and Cys474 in keratin 5; Cys432 in keratin 9; Cys40, Cys18, and Cys389 in keratin 14; and Cys49 and Cys497 in keratin 1), as well as microfilament and microtubule filament proteins, such as tubulin and actins.

## Discussion

### Summary of evidence

Deamidation was found to be the major age-related PTM and large numbers of asparagine and glutamine sites were found to be significantly more deamidated in the water-insoluble proteins of aged lenses. Some crystallins from older lenses contained sites were almost totally deamidated, including βA1/A3-, βA4-, γS-, and γD-crystallins. Some sites showed increased levels of deamidation in ARNC cataractous lenses. No significant difference in the extent of Gln92 and Gln170 deamidation of αΑ-crystalin in D-isoAsp58 between cataract and age-matched normal lenses.

There was a significant increase in the amount of αΑ-crystalin in D-isoAsp58 present at all ages. This was most evident during the early years, when age-related cataract is first observed.

An increase in the extent of Trp oxidation was apparent in cataract lenses when compared to aged normal lenses. No phosphorylated Ser residues were detected in αB-crystallin from the cataract lenses, whereas up to four were observed in the young and aged normal lens samples. Οne new βB1-crystallin truncation was identified in both the aged normal and cataract lenses. No Arg+55 Da modifications were detected in the cataract lenses. Twenty-two previously unreported proteins were identified in normal aged and 29 in cataract lenses. Phakinin, protein DJ-1 and phosphoglycerate kinase 1, where both cataract and aged normal lenses had greater relative abundance comparing to foetal lenses. Several proteins had a high abundance in the foetal lens group but a much reduced level in both adult lens groups, including betaine– homocysteine S-methyltransferase 1, bA2-crystallin, bB3-crystallin, gC-crystallin, glycer-aldehyde-3-phosphate dehydrogenase, per oxiredoxin-6 and vimentin.

There was evidence of age-related shift from intra- to intermolecular disulfide formation in human and LEGSKO Mouse Lens Crystallins. Four types of cysteine residues were identified based on their susceptibility to oxidation with age and as a function of presence and severity of cataract graded from II to V. The first includes cysteine residues that are mildly oxidized with age and not further oxidized in cataracts; the second includes cysteine residues with mild oxidation in old normal lenses but massive increase in cataracts regardless of cataract severity; the third includes cysteine residues with oxidation in old normal lenses and further increase in cataract lenses, and the fourth includes cysteine residues that are barely oxidized with age but oxidized in cataracts, and also associated with cataract severity.

Enzymes with oxidized cysteine at critical sites included GAPDH, glutathione synthase, aldehyde dehydrogenase, sorbitol dehydrogenase, and PARK7. They stated that extensive oxidation was also present in lens-specific intermediate filament proteins, such as BFSP1 and BFSP12, vimentin, cytokeratins, the majority of which were positively associated with degrees of cataract severity, as well as microfilament and microtubule filament proteins, such as tubulin and actins.

### Limitations

The present systematic review should be interpreted within the context of its strengths and limitations.

Outcome level: The main limitation of this systematic review is that the outcome of interest is not the same across studies.

Study and review level: Our study has some limitations. The quality of the included studies showed a low risk of bias although four studies included a small number of subjects/lenses. However, all studies used proteomic analysis that has a tremendous favorable potential for detecting candidate proteins and, so, facilitating the development of precision medicine for diseases such as age-related cataract.

## Conclusion

The results of Hooi et al. showed that in the case of Gln92 and Gln170, the degree of deamidation did not differ significantly between cataract and age-matched normal lenses and this may suggest that these particular modifications may contribute to age-related changes to the physical properties of the human lens, but not to cataractogenesis. Their result that there is a consistently greater extent of deamidation of Asn 76 in γS-crystallin in all cataract lenses compared with their age-matched controls, suggests that this modification, along with others, such as deamidation at Asn 14 and Gln16, could be associated with the development of age-related cataract in human. In addition to this, there was a consistently greater formation of D-isoAsp in αA crystalline from cataract lenses compared with age-matched normal lenses, suggesting that it could as well be associated with the development of age-related cataract in human. Such a dominant age-related modification may contribute to the denaturation of a crystallin, and its ability to act as a chaperone.

Based on the results from their proteome analysis of human foetal, aged and advanced nuclear cataract lenses, Hains and Truscott suggested there are no major differences between aged normal and advanced cataract lenses in terms of proteins present.

The results of their other study showed that the level of Trp oxidation appeared to increase in cataract lenses when these were compared to age-matched normal lenses and this may indicate that oxidation of Trp may be associated with nuclear cataract formation in man.

Deamidation appears to be important for protein denaturation in the aging lens and it is possible that the few sites they observed to be differentially deamidated in cataractous lenses may be sufficient to cause opacification.

The results of Wilmarth et al. show that large numbers of asparagine and glutamine sites were significantly more deamidated in the water-insoluble proteins of aged lenses, and since cataract can be thought of as a crystallin insolubility disease, modifications exclusively associated with water-insoluble proteins may have implications in cataract formation.

Finally, the results of Wang et al. and fan et al. support the important role of oxidant stress in age-related cataract formation, and in particular, the importance of GSH and that pharmacological GSH mimetics are probably needed to protect the aging lens from oxidation.

## Data Availability

Not applicable

## Abbreviations

(GSH): Glutathione
(SH): Sulfhydryl
(PRISMA): Preferred Reporting Items for Systematic Reviews and Meta-Analyses (WI-US) Water-insoluble-urea soluble
(WI-UI): Water-insoluble-urea insoluble
(ARNC): Age-related nuclear cataract
(NEM): N-ethylmaleimide
(CM): Carboxymethyl
(HMW): High Molecular Weight
(Gln): Glutamine
(FABP5): Fatty acid-binding protein
(CRYAB): Alpha-crystallin B chain
(BHMT1): Betaine-homocysteine S-methyltransferase-1
(RLSC): Regenerative lenses with secondary cataract
(CC): Congenital cataract
(ARC): Age-related cataract
(VEGF): Vascular endothelial growth factor
(hγD): human γD-crystallin

## Aknowledgment

We thank Anastasios Varypatis for helping us design the graphical illustration.

## References

1. Crystalline Lens and Cataract by Joah F. Aliancy and Nick Mamalis – Webvision [Internet]. [cited 2018 Sep 27]. Available from: https://webvision.med.utah.edu/book/part-xvi-anterior-segment/crystalline-lens-and-cataract/

2. Pascolini D, Mariotti SP. Global estimates of visual impairment: 2010. Br J Ophthalmol [Internet]. 2012 May [cited 2018 Oct 6];96(5):614–8. Available from: http://www.ncbi.nlm.nih.gov/pubmed/22133988

3. Zetterberg M, Celojevic D. Gender and Cataract – The Role of Estrogen. Curr Eye Res [Internet]. 2015 Feb 2 [cited 2018 Oct 6];40(2):176–90. Available from: http://www.ncbi.nlm.nih.gov/pubmed/24987869

4. Resnikoff S, Pascolini D, Etya’ale D, Kocur I, Pararajasegaram R, Pokharel GP, et al. Global data on visual impairment in the year 2002. Bull World Health Organ [Internet]. 2004 Nov [cited 2018 Oct 6];82(11):844–51. Available from: http://www.ncbi.nlm.nih.gov/pubmed/15640920

5. Prokofyeva E, Wegener A, Zrenner E. Cataract prevalence and prevention in Europe: a literature review. Acta Ophthalmol [Internet]. 2013 Aug 1 [cited 2018 Oct 6];91(5):395–405. Available from: http://doi.wiley.com/10.1111/j.1755-3768.2012.02444.x

6. Ottonello S, Foroni C, Carta A, Petrucco S, Maraini G. Oxidative Stress and Age-Related Cataract. Ophthalmologica [Internet]. 2000 [cited 2018 Oct 14];214(1):78–85. Available from: http://www.ncbi.nlm.nih.gov/pubmed/10657746

7. Truscott RJW. Age-related nuclear cataract—oxidation is the key. Exp Eye Res [Internet]. 2005 May [cited 2018 Oct 7];80(5):709–25. Available from: http://www.ncbi.nlm.nih.gov/pubmed/15862178

8. anatomy & physiology of lens [Internet]. [cited 2018 Oct 7]. Available from: <https://www.slideshare.net/rakeshjaiswal10/anatomy-physiology-of-lens>

9. Hoehenwarter W, Klose J, Jungblut PR. Eye lens proteomics. Amino Acids [Internet]. 2006 Jun 4 [cited 2018 Oct 23];30(4):369–89. Available from: http://www.ncbi.nlm.nih.gov/pubmed/16583312

10. Psatha K, Kollipara L, Voutyraki C, Divanach P, Sickmann A, Rassidakis GZ, et al. Deciphering lymphoma pathogenesis via state-of-the-art mass spectrometry-based quantitative proteomics. J Chromatogr B [Internet]. 2017 Mar 15 [cited 2018 Dec 9];1047:2–14. Available from: <https://www.sciencedirect.com/science/article/pii/S1570023216306432>

11. Association for Research in Vision and Ophthalmology. Y, Duncan MK, David LL. Investigative ophthalmology & visual science. [Internet]. Vol. 43, Investigative Ophthalmology & Visual Science. [Association for Research in Vision and Ophthalmology, etc.]; 2002 [cited 2018 Oct 23]. 205–215 p. Available from: https://iovs.arvojournals.org/article.aspx?articleid=2123357

12. Ahmad MT, Zhang P, Dufresne C, Ferrucci L, Semba RD. The Human Eye Proteome Project: Updates on an Emerging Proteome. Proteomics [Internet]. 2018 Mar [cited 2018 Oct 23];18(5–6):1700394. Available from: http://www.ncbi.nlm.nih.gov/pubmed/29356342

13. PRISMA [Internet]. [cited 2018 Oct 24]. Available from: http://www.prisma-statement.org/PRISMAStatement/Checklist

14. Wilmarth PA, Tanner S, Dasari S, Nagalla SR, Riviere MA, Bafna V, et al. Age-Related Changes in Human Crystallins Determined from Comparative Analysis of Post-translational Modifications in Young and Aged Lens: Does Deamidation Contribute to Crystallin Insolubility? J Proteome Res [Internet]. 2006 Oct [cited 2019 May 14];5(10):2554–66. Available from: http://www.ncbi.nlm.nih.gov/pubmed/17022627

15. Hains PG, Truscott RJW. Post-Translational Modifications in the Nuclear Region of Young, Aged, and Cataract Human Lenses. J Proteome Res [Internet]. 2007 Oct [cited 2019 May 15];6(10):3935–43. Available from: http://www.ncbi.nlm.nih.gov/pubmed/17824632

16. Hains PG, Truscott RJW. Proteome analysis of human foetal, aged and advanced nuclear cataract lenses. PROTEOMICS - Clin Appl [Internet]. 2008 Dec [cited 2019 May 27];2(12):1611–9. Available from: http://www.ncbi.nlm.nih.gov/pubmed/21136811

17. Hains PG, Truscott RJW. Age-Dependent Deamidation of Lifelong Proteins in the Human Lens. Investig Opthalmology Vis Sci [Internet]. 2010 Jun 1 [cited 2019 May 23];51(6):3107. Available from: http://iovs.arvojournals.org/article.aspx?doi=10.1167/iovs.09-4308

18. Hooi MYS, Raftery MJ, Truscott RJW. Racemization of Two Proteins over Our Lifespan: Deamidation of Asparagine 76 in γS Crystallin Is Greater in Cataract than in Normal Lenses across the Age Range. Investig Opthalmology Vis Sci [Internet]. 2012 Jun 14 [cited 2019 May 23];53(7):3554. Available from: http://www.ncbi.nlm.nih.gov/pubmed/22531704

19. Hooi MYS, Raftery MJ, Truscott RJW. Age-dependent deamidation of glutamine residues in human γS crystallin: Deamidation and unstructured regions. Protein Sci [Internet]. 2012 Jul [cited 2019 May 14];21(7):1074–9. Available from: http://www.ncbi.nlm.nih.gov/pubmed/22593035

20. Hooi MYS, Raftery MJ, Truscott RJW. Accelerated aging of Asp 58 in αA crystallin and human cataract formation. Exp Eye Res [Internet]. 2013 Jan [cited 2019 May 23];106:34–9. Available from: https://linkinghub.elsevier.com/retrieve/pii/S0014483512003181

21. Fan X, Zhou S, Wang B, Hom G, Guo M, Li B, et al. Evidence of Highly Conserved β-Crystallin Disulfidome that Can be Mimicked by In Vitro Oxidation in Age-related Human Cataract and Glutathione Depleted Mouse Lens. Mol Cell Proteomics [Internet]. 2015 Dec [cited 2019 May 15];14(12):3211–23. Available from: http://www.ncbi.nlm.nih.gov/pubmed/26453637

22. Wang B, Hom G, Zhou S, Guo M, Li B, Yang J, et al. The oxidized thiol proteome in aging and cataractous mouse and human lens revealed by ICAT labeling. Aging Cell [Internet]. 2017 Apr [cited 2019 May 15];16(2):244–61. Available from: http://www.ncbi.nlm.nih.gov/pubmed/28177569

